# Lifestyle acquired immunity, decentralized intelligent infrastructures and revised healthcare expenditures may limit pandemic catastrophe: a lesson from COVID-19

**DOI:** 10.1101/2020.05.23.20111104

**Authors:** Asif Ahmed, Tasnima Haque, Mohammad Mahmudur Rahman

## Abstract

Human race has often faced pandemic with substantial number of fatalities. As COVID-19 pandemic reached and endured in every corner on earth, countries with moderate to strong healthcare support and expenditure seemed to struggle in containing disease transmission and casualties. COVID-19 affected countries have variability in demographic, socioeconomic and life style health indicators. At this context it is important to find out at what extent these parametric variations are actually modulating disease outcomes. To answer this, we have selected demographic, socioeconomic and health indicators e.g. population density, percentage of urban population, median age, health expenditure per capita, obesity, diabetes prevalence, alcohol intake, tobacco use, case fatality of non communicable diseases (NCDs) as independent variables. Countries were grouped according to these variables and influence on dependent variables e.g. COVID-19 test positive, case fatality and case recovery rates were statistically analyzed. The results suggest that countries with variable median age has significantly different outcome on test positive rate (*P*<0.01). Both median age (*P*=0.0397) and health expenditure per capita (*P*=0.0041) has positive relation with case recovery. Increasing number of test per 100K population showed positive and negative relation with number of positives per 100K population (*P*=0.0001) and percentage of test positives (*P*<0.0001) respectively. Alcohol intake per capita in liter (*P*=0.0046), diabetes prevalence (*P*=0.0389) and NCDs mortalities (*P*=0.0477) also showed statistical relation with case fatality rate. Further analysis revealed that countries with high healthcare expenditure along with high median age and increased urban population showed more case fatality but also had better recovery rate. Investment in health sector alone is insufficient in controlling pandemic severity. Intelligent and sustainable healthcare both in urban and rural settings and healthy lifestyle acquired immunity may reduce disease transmission and comorbidity induced fatalities respectively.

## 1 Introduction

Rapid demographic changes are being observed in most of the region and countries since the middle of the last century. Increased population density, urban population and life expectancy are noticeable examples of such changes (1, 2). The key objective of this transition is to improve socioeconomic level of a country’s population. Socioeconomic development influence population health status through regulation of environment, lifestyle, and healthcare systems (3). Socioeconomic variables viz. population density, gross national income (GNI) per capita and health expenditures per capita play important role to achieve sustainable development (4, 5). The health and allied policies of governments are important and perhaps the most critical aspect to ensure facilities as well as management of population (6).

Last few decades many governments are spending in health sectors to improve the healthcare systems, treatments, research and development of new drugs and vaccines and technologies for preventing and controlling of diseases (7-12). A lion’s share of this fund is being invested to prevent and treat diseases of communicable and non-communicable nature. Amount spent on health is mostly depended on GNI at purchasing power parity (PPP) (5, 13, 14) and the outcome is visible as reduced child mortality, increase in median age and life expectancy at birth (15-18). Therefore, life expectancy, median age and percentages of urban population have become the key indicators in human development indexes (19-21). Top listed economies by GNI (PPP) are conventionally ahead in technology, research and training (22).

Chronic respiratory diseases are responsible for almost four million premature deaths globally (23). Respiratory infections are usually worsened through population density, human behavior, insufficient public health safety, genomic mutations of microbes, extraneous usage and developing resistance to antibiotics. Lack of global coordination to prevent infectious disease outbreak and pandemic is partly due to weak policies, management and expenditures in autocratic regimes pushing global health security at risk (24). In addition, developed countries created facilities, readiness and prevention from several life-threatening diseases without any divergences in rural and urban populations (24). However, these sustainably developed countries are facing devastating disasters in new Corona Virus Disease-19 (COVID-19) pandemic (25). COVID-19 emerged in December 2019 in Wuhan, China and undergoing with numbers of morbidity and mortality worldwide (26). Till date as of May 22, 2020, there are more than 5.1 million positive cases and more than 0.3 million deaths (27).

COVID-19 pandemic is caused by a positive-sense single stranded RNA (+ssRNA) virus named SARS-CoV-2 which belongs to corona virus family. Family of these viruses are capable of introducing human sickness (26) with an incubation periods ranging from 2 to 14 days to develop symptoms (28). SARS-CoV-2 is mostly transmitted between persons via respiratory droplets, coughs, sneezes and fomites (2). COVID-19 patients could be asymptomatic or develop flu-like symptoms with fever, dry cough, tiredness and shortness of breath. Intensive cares with ventilation, with symptom based therapies are needed for critical patients (2). The World Health Organization (WHO) declared COVID-19 outbreak as Public Health Emergency of International Concern (PHEIC) on 30 January and pandemic on 11 March 2020 (29).

Clinical reports confirmed that non-communicable disease (NCDs) including diabetes, heart disease, hypertension, respiratory disease (COPD/bronchial asthma), cancer, predominantly amongst the aged individuals, upsurge the susceptibility to COVID-19 (30). Surprisingly, countries with developed facilities to manage NCDs are struggling in COVID-19 pandemic. Healthcare personnel are also being infected in all countries regardless of country’s economic status and demographic characters (24, 31). Several disease susceptibility pattern and prediction were also made to know trends of infections, death and recovery patterns (6, 32) as well as factors that influenced the transmissions and fatalities (6). Associations of different environmental factors have also been investigated (6, 33, 34). COVID-19 requires sufficient public attention, and need to reprioritize the financial involvement in appropriate segments of health sector to confirm inclusive responses. Highly affected countries have taken numerous tactics of financial distribution, depending on their capabilities, structures and regulatory systems (35). In-depth investigations need to be conducted on the association of infection pattern, fatalities and recovery in combination with population density, median age, percentage of urban population, GDP per capita and health expenditure per capita along with lifestyle and health indicators.

In this study, publicly available demographic, socioeconomic and health indicators data of COVID-19 affected countries have been analyzed to extrapolate influences of categorical independent variables on disease outcome. The main research question was at what degree these variables have significant impact on the dependent variables including test positives rates, case fatality rates and case recovery rates. These findings along with other studies of similar nature might help to strengthen preparedness to face any yet-to-come contagious pandemic in near future. The study findings also highlight the importance of implementing intelligent healthcare system both in urban and rural areas coupled with healthy lifestyle that boost population immunity. Refocusing healthcare investment on poorly addressed sectors should be prioritized to minimize life and economic loss at global scale.

## 2 Methods

### 2.1 Data characteristics

The study was designed to use available secondary data for all variables. The COVID-19 data of total positive cases, total death cases, total recovered cases and total tests cases data were obtained from Worldometers.info (27). Socioeconomic and demographic data including total population, population density, median age, urban population percentages, male and female ratio and financial information together with gross domestic product (GDP) in USD, gross national income (GNI) per capita (purchasing power parity, PPP) in USD, health expenditure (% of GDP) in USD were attained from the databank of World Bank (36). However, health expenditure per capita data of Hong Kong was obtained from the website of Department of Health of the Government of the Hong Kong Special Administrative Region (37). In addition, GNI per capita (PPP) of Djibouti was obtained from the database of International Monetary Fund (38). Lifestyle and health indicator data including prevalence of obesity (39), prevalence of insufficient physical activity (40), prevalence of diabetes (41), NCD mortality rate per 100K population (42), alcohol consumption per capita per liter per year (43) and tobacco use percentages of male (44) were obtained from WHO and World Bank.

### 2.2 Inclusion and exclusion criteria

The top 91 countries were selected decisively whose total infection cases crossed 1000 on May 09, 2020. For any particular parameter, some country data were unavailable; therefore, these countries were excluded in the relevant analysis, however, other data from these countries were used in respective analysis.

### 2.3 Variables

The current study was conducted with several variables. Variables related to COVID-19, viz. total positive cases, total death cases, total recovered cases, total number of tests were used. Percentages of test positives were calculated accordingly. Case fatality and case recovered rate were calculated against total positive cases. Percentage of test positives, case fatality and case recovered rate were used as dependent variables instead of total positive cases, death cases and recovery cases as we assumed due to the socioeconomic condition a number of countries did not go through mass diagnosis strategies instead, they conducted diagnosis when COVID-19 related symptoms appeared. Population density, median age, percentages of urban population, GNI per capita (PPP) in USD and health expenditures per capita in USD was used as socioeconomic and demographic independent variables. Lifestyle and health related independent variables viz. prevalence of obesity, insufficient physical activity, diabetes and any kind of tobacco use by male and female were obtained as percentages. Alcohol consumption per capita per liter variables were the average number of liters consumed per year were considered. In addition, NCDs mortality rate per 100K population were also taken as another independent variables. Tests numbers of countries were not uniform and depend on several factors including socioeconomic status and government initiatives. Thus, we calculated the number of tests per 100K populations as well as positive case per 100K populations. Tests per 100K population were used as independent variables and analyzed against test positive rates, case fatalities and case recovered rate along with positive cases per 100k as dependent variables. Linear regression analysis was performed with ungrouped independent variables with dependent variables as one to one analysis fashion. Later group wise distribution of independent variables was analyzed with dependent variables to find the association.

### 2.4 Independent variables processing and grouping

Independent variables except GNI per capita (PPP) were grouped for getting trends and scenarios in relation with dependent variables (**Table 1**). GNI per capita (PPP) in USD data were grouped into categories according to the World Bank (45). Grouped independent variables were analyzed against dependent variables as mean ± SEM. Cross analysis of each independent variable were also performed to get in depth information and narrated in discussion. Cross connection analysis results were tabulated and placed in supplementary.

**Table 1:** Characteristics of grouped independent variables.

| **Variables** | **Groups description** | **Numbers of Country** | **Unavailable data (no of country)** |
| --- | --- | --- | --- |
| **Population density Km<sup>2</sup>** | Up to 100 | 51 (56.04%) | 0 |
|  | 100+ to 250 | 26 (28.57%) |  |
|  | 250+ to 500 | 8 (8.79%) |  |
|  | 500+ | 6 (6.59%) |  |
| **Per capita GNI (PPP) in USD\*** | Up to 1026 | 0 | 0 |
|  | 1026+ to 3995 | 4 (3.40%) |  |
|  | 3995+ to 12375 | 19 (20.88%) |  |
|  | 12375+ | 68 (74.73%) |  |
| **Urban population, %** | Up to 50% | 12 (13.19%) | 0 |
|  | 50%+ to 75% | 38 (41.76%) |  |
|  | 75%+ | 41 (45.05%) |  |
| **Median age, years** | Up to 20 | 6 (6.59%) | 0 |
|  | 20+ to 30 | 21 (23.08%) |  |
|  | 30+ to 40 | 30 (32.97%) |  |
|  | 40+ | 34 (37.36%) |  |
| **Per Capita Health Expenditure in USD** | U to 100 | 12 (13.19%) | 0 |
|  | 100+ to 500 | 28 (30.77%) |  |
|  | 500+ to 2000 | 26 (28.57%) |  |
|  | 2000+ | 25 (27.47%) |  |
| **Prevalence of obesity, %** | Up to 15% | 19 (20.88%) | 3 (3.30%) |
|  | 15%+ to 20% | 41 (45.05%) |  |
|  | 20%+ | 28 (30.77%) |  |
| **Prevalence of insufficient physical activities, %** | Below 20% | 9 (9.89%) | 14 (15.38%) |
|  | 20% to 30% | 22 (24.18%) |  |
|  | 30%+ to 40% | 35 (38.46%) |  |
|  | 40%+ | 11 (12.09%) |  |
| **NCD mortality per 100K** | Up to 300 | 7 (7.69%) | 5 (5.49%) |
|  | 300+ to 400 | 30 (32.97%) |  |
|  | 400+ to 500 | 22 (24.18%) |  |
|  | 500+ to 600 | 14 (15.38%) |  |
|  | 600+ to 700 | 11 (12.09%) |  |
|  | 700+ | 12 (13.19%) |  |
| **Prevalence of diabetes, %** | Up to 5% | 16 (17.58%) | 1 (1.10%) |
|  | 5%+ to 10% | 56 (61.54%) |  |
|  | 10% to 15% | 8 (8.79%) |  |
|  | 15%+ | 8 (8.79%) |  |
| **Alcohol consumptions per capita in Liters per year** | Up to 5 L | 32 (35.16%) | 2 (2.20%) |
|  | 5+ L to 10 L | 35 (38.46%) |  |
|  | 10+ | 16 (17.58%) |  |
| **Any kind of tobacco used by male, % of population** | Up to 25% | 24 (26.37%) | 2 (2.20%) |
|  | 25% to 40% | 29 (31.87%) |  |
|  | 40%+ | 19 (20.88%) |  |

*[Groups were formed according to the distribution. Grouping of Per capita GNI (PPP) was done according to the World Bank classification by income level]*

### 2.5 Statistical analysis

One to one regression analysis of ungrouped data was performed in SPSS version 26. GraphPad Prism version 6 was used to generate graphs. One- or two- way ANOVA were performed as required along with individual group to group statistical variation analysis. All statistical significance was measured at significant value <0.05. All the final graphs generated in GraphPad Prism were combined using Inkscape version 0.92 graphics software.

## 3 Results

COVID-19 pandemic is showing uneven epidemiological and clinical trend as it is spreading to countries with climatic, socio economic, lifestyle and demographic variation around the globe. In addition to mutation induced genomic variations in the virus, these factors might have substantial influence in key outcomes like rate of infection, case fatality and case recovery. This study was conducted to find the possible link between rate of COVID-19 infections, fatalities and recovery with socioeconomic, demographic, lifestyles and health indicators.

Associations of dependent variables with non-grouped independent variables were measured with linear regression and results were shown in **Table 2**. One to one regression analysis showed that, median age (*P*=0.005), any kind of tobacco use by male (*P*=0.004) and female (*P*=0.02) were significantly linked with test positive rate. Case fatality rates were associated and significantly predicted by male/female ration of population (*P*=0.033), median age (*P*=0.005), health expenditure per capita in USD (*P*=0.008), NCD mortalities per 100K (*P*=0.003), alcohol consumptions per capita in liter (*P*=0.001) and tobacco used by female (*P*=0.007). Moreover, case recovered rates were also significantly related with median age (*P*=0.004), health expenditure per capita in USD (*P*=0.018), and NCD mortalities per 100K (*P*=0.038).

**Table 2:** One to one linear regression analysis using ungrouped independent variables.

|  | Test Positive Rate |  |  |  | Case Fatality Rate |  |  |  | Case Recovered Rate |  |  |  |
| --- | --- | --- | --- | --- | --- | --- | --- | --- | --- | --- | --- | --- |
|  | R<sup>2</sup> | F | Regressi<br>on<br>Coefficie<br>nts (β) | p | R<sup>2</sup> | F | Regressi<br>on<br>Coefficie<br>nts (β) | p | R<sup>2</sup> | F | Regressi<br>on<br>Coefficie<br>nts (β) | p |
| Male/Female ratio | 0.001 | 0.059 | -0.027 | 0.808 | 0.051 | 4.694 | -0.225 | 0.033 | 0.035 | 3.109 | -0.186 | 0.081 |
| Population Density/km<sup>2</sup> | 0.003 | 0.29 | -0.058 | 0.592 | 0.029 | 2.667 | -0.171 | 0.106 | 0 | 0.026 | 0.017 | 0.873 |
| Urban Population, % | 0.001 | 0.067 | -0.028 | 0.797 | 0.011 | 0.994 | 0.105 | 0.321 | 0.035 | 3.137 | 0.187 | 0.08 |
| Median Age, Years | 0.091 | 8.478 | -0.301 | 0.005 | 0.086 | 8.393 | 0.294 | 0.005 | 0.092 | 8.787 | 0.303 | 0.004 |
| PC GNI (PPP) in USD | 0.035 | 3.063 | -0.186 | 0.084 | 0.003 | 0.254 | 0.053 | 0.616 | 0.024 | 2.133 | 0.155 | 0.148 |
| PC Health Expenditure in USD | 0.013 | 1.123 | -0.114 | 0.292 | 0.076 | 7.325 | 0.276 | 0.008 | 0.062 | 5.775 | 0.249 | 0.018 |
| Obesity prevalence, % | 0 | 0.032 | 0.020 | 0.859 | 0.009 | 0.793 | 0.096 | 0.376 | 0.006 | 0.501 | 0.077 | 0.481 |
| Insufficient Physical activity Prevalence, % | 0 | 0.008 | -0.011 | 0.929 | 0.003 | 0.234 | 0.056 | 0.63 | 0.018 | 1.341 | -0.133 | 0.251 |
| NCD Mortality per 100K | 0 | 0.005 | 0.008 | 0.945 | 0.099 | 9.236 | -0.315 | 0.003 | 0.051 | 4.453 | -0.226 | 0.038 |
| Diabetes prevalence, % | 0.005 | 0.425 | 0.071 | 0.516 | 0.008 | 0.666 | -0.087 | 0.417 | 0.013 | 1.162 | -0.115 | 0.284 |
| Alcohol consumptions per capita per L | 0.042 | 3.659 | -0.205 | 0.059 | 0.11 | 10.767 | 0.332 | 0.001 | 0.04 | 3.554 | 0.199 | 0.063 |
| Tobacco used by male % | 0.097 | 8.95 | -0.312 | 0.004 | 0 | 0.042 | -0.022 | 0.838 | 0 | 0.001 | -0.004 | 0.073 |
| Tobacco used by female % | 0.064 | 5.645 | -0.252 | 0.02 | 0.081 | 7.678 | 0.285 | 0.007 | 0.017 | 1.522 | 0.132 | 0.221 |

*[Linear regression analysis was conducted as one to one analysis with dependent and independent variable. Significant value was calculated as P=<0.05 and mentioned in italic in the table. ]*

The three dependent variables of the present study covering 91 countries have mean percentages of positive cases per test, case fatality and case recovery of 9.94% ± 1.25, 4.26% ± 0.38 and 44.98% ± 2.80 respectively. The goal was to find any significant differences among these variables when countries were grouped according to different socioeconomic, demographic, life style and health determinants.

### 3.1 Effect of socioeconomic and demographic factors on percentages of test positives

Population density, percentage of urban population, median age and health expenditure per capita were used as independent variables. These variables were grouped as mentioned in Table 1. Data of total tests performed were not available for four countries (Cameroon, China, Guinea and Sudan), therefore 87 countries were included in this section. Results showed that, countries with low population density (below 100 people per km^2^) had high percentage of test positives (Figure 1A, left panel). However, countries in other population density groups had lower test positive rates with minimum variability among them. Percent of test positive were slightly lower in countries with high urban density (75% above) compared to other groups (Figure 1B, left panel). Countries with increasing median age showed statistically significant decrease in percent test positives (Figure 1C, left panel). The relation between percentage of test positive and health expenditure per capita showed no significance although countries spending higher had lower outcome (Figure 1D, left panel).

**Figure 1:**
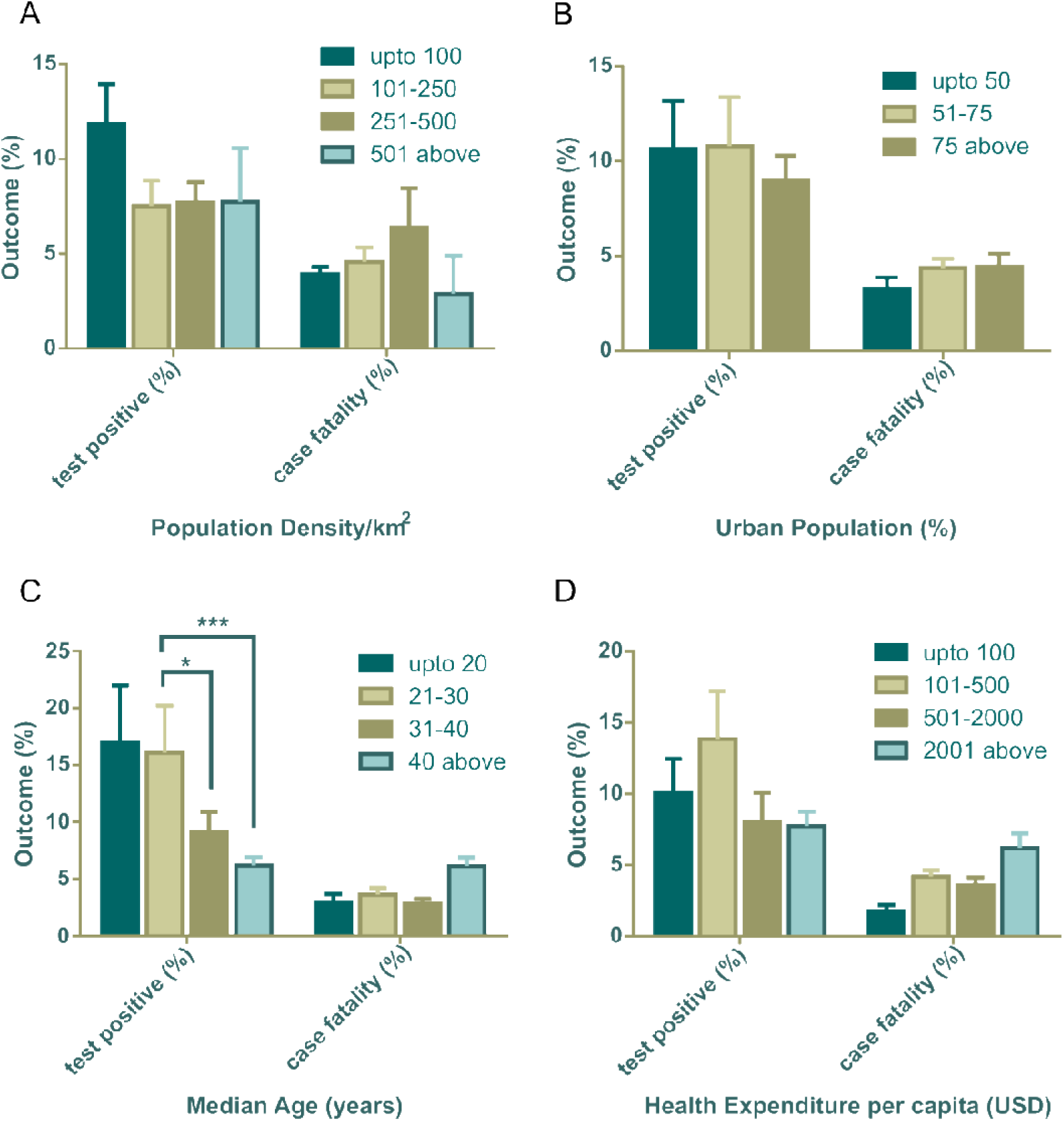
Test positive (%) and case fatality (%) distribution among countries categorized with population density (A), urban population (B), median age (C) and health expenditure per capita (D). All outcomes (%) are presented as mean ± SEM. In Fig. 1(C), mean test positive (%) of countries with 21-30 years of median age is statistically significant against 31-40 years of median age (\**P*=0.0180) and median age over 40 years (\*\*\**P*=0.0001) by Tukey’s multiple comparison test. Two-way ANOVA of rest of the data are statistically non-significant.

### 3.2 Effect of socioeconomic and demographic factors on case fatality rate

Percent case fatalities and its association with population density was uneven and was not statistically significant (Figure 1A, right panel), however, countries with higher urban population showed higher case fatalities (Figure 1B, right panel). Although, percentage of test positives had significant association with median age, case fatalities did not show the trend (Figure 1C, right panel). But, countries with highest number of older population (median age over 40 years) confronted higher fatality rate compared to other three groups. Similarly, case mortality was found to be high in the countries where government disburses more in health sector per capita (Figure 1D, right panel).

### 3.3 Effect of socioeconomic and demographic factors on case recovery rate

The association of socioeconomic and demographic determinants with case recovery rate was shown in Figure 2. Population density did not show any connection with case recovered rate (Figure 2A). Similar to the case fatalities trend, countries with high urban population showed higher recovery rate than the countries with lower dense city population (Figure 2B). As like case fatalities, countries with increasing median age showed increased recovery rate (Figure 2C) and variances of mean values were statistically significant by one-way ANOVA (\**P*=0.0397). Figure 2D signified that, countries spending more in healthcare systems per capita, had more recovery rates from COVID-19 (\*\**P*=0.0041) (Figure 2D). Due to unavailability of total recovered cases information of the Netherlands and the United Kingdom, they were not included in this segment of analysis.

**Figure 2:**
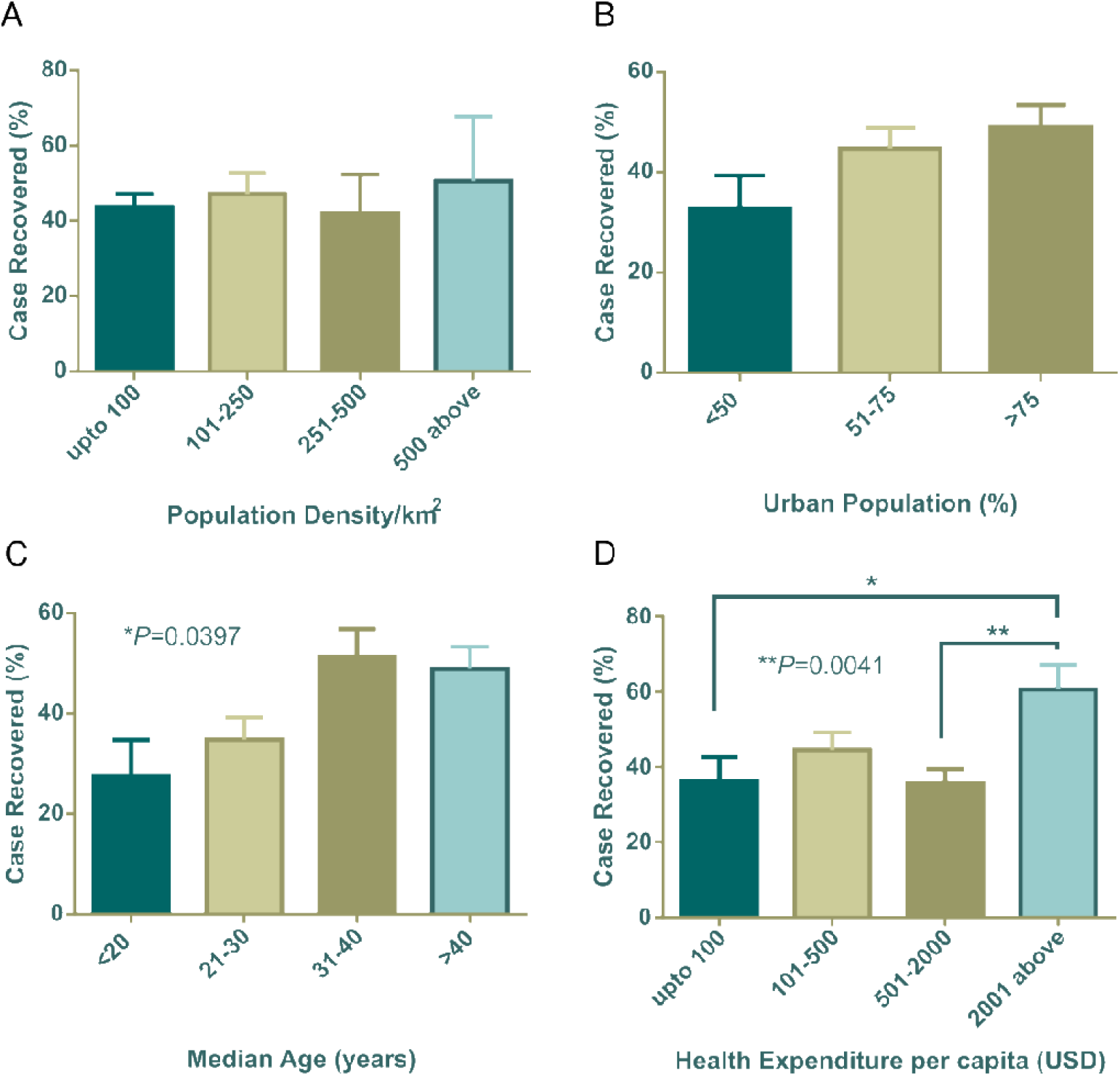
Case recovery (%) distribution among countries categorized with population density (A), urban population (B), median age (C) and health expenditure per capita (D). In Fig. 2(C) and 2(D), differences among means are statistically significant using one way ANOVA. Rest of the data shows no significant variation. Data presented are mean ± SEM.

### 3.4 Effect of number of tests on rate of test positives, fatalities and recovery

All countries included in the study varied in COVID-19 diagnosis capacity due to social, economic and political reasons. Thus, we wanted to further know whether the number of tests performed per 100K population had any significant effect on mean positive cases, percentage of case positives, case fatality and case recovery. To do this analysis, we calculated country specific number of positive cases/100K population and tests performed per 100K population from data of the number of total test (27), total positive cases (27) and the total population of the country (36).

Association of COVID-19 test numbers per 100K population was presented in Figure 3. Positive cases per 100K populations were significantly (\*\*\*\**P*<0.001) boosted with the increased number of tests performed per 100K populations (Figure 3A), in contrast, percentage of test positives were high in lowest COVID-19 tests per 100K populations (\*\*\*\**P*<0.0001) (Figure 3B). Case fatality did not change significantly among countries grouped into increasing test numbers (Figure 3C). Higher case recovery rates were observed in the groups of countries where higher number of tests were performed (Figure 3D). Cameroon, China, Guinea and Sudan were out of this analysis due to unavailability of tests data from these countries.

**Figure 3:**
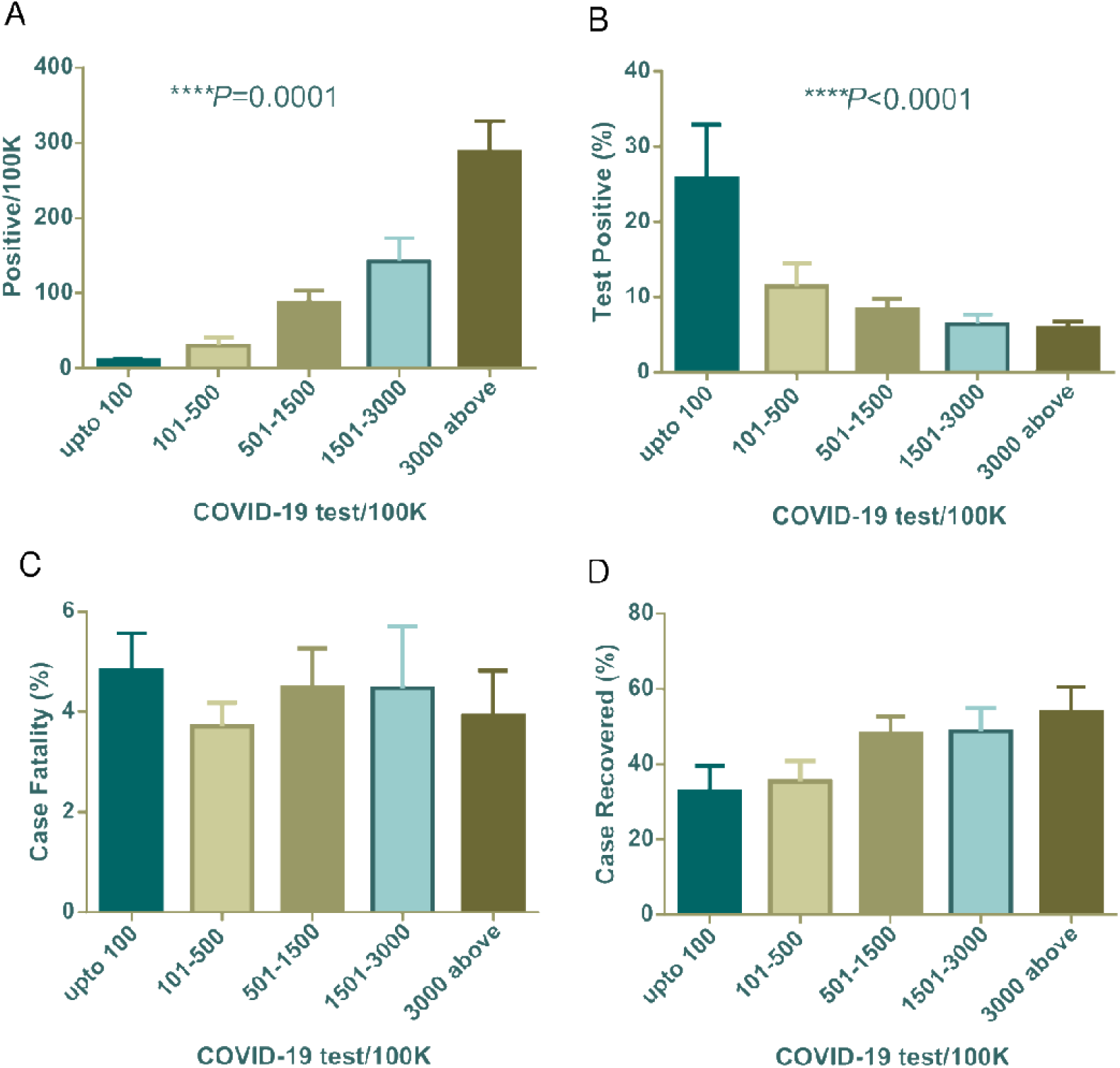
Positives/100K, test positive (%), case fatality (%) and case recovery (%) data distribution among countries grouped with COVID-19 test/100K population. All are mean ± SEM. In one-way ANOVA variance among column means are statistically significant for Fig. 3(A) and 3(B).

### 3.5 Lifestyle and health indicators

Individual, community and/or social lifestyle and practices can influence transmissions of viruses as well as death and recovery rates. Study showed that health indicators like prevalence of diabetes and other NCDs also influence the number of death and recovery (46). Therefore, we considered prevalence of obesity, insufficient physical activities, diabetes, NCDs mortality per 100K population, consumption of alcohol per capita per liter per year as well as percent of general tobacco uses in male populations as independent variables.

#### 3.5.1 Lifestyle and health indicators on case fatalities of COVID-19

In our investigation, countries with high obesity prevalence were witnessing more COVID-19 related death (Figure 4A) and correspondingly countries where people were less physically active also beholder of increased death rate (Figure 4B). Countries with high alcohol consumption were significantly (*P*=0.0046) prone to death (Figure 4C). However, interestingly tobacco use pattern showed different scenarios compared to alcohol consumption. Higher percentages of any kind of tobacco used in males showed less case fatalities compared to other groups of countries (Figure 4 D). With further surprise countries with low diabetes prevalence showed significantly (*P*=0.0389) higher death rate (Figure 4E). NCD mortality per 100K population showed negative relation with COVID-19 case fatality rate (*P*=0.0477) where countries with high NCD mortality experienced less COVID-19 case fatality (Figure 4F).

**Figure 4:**
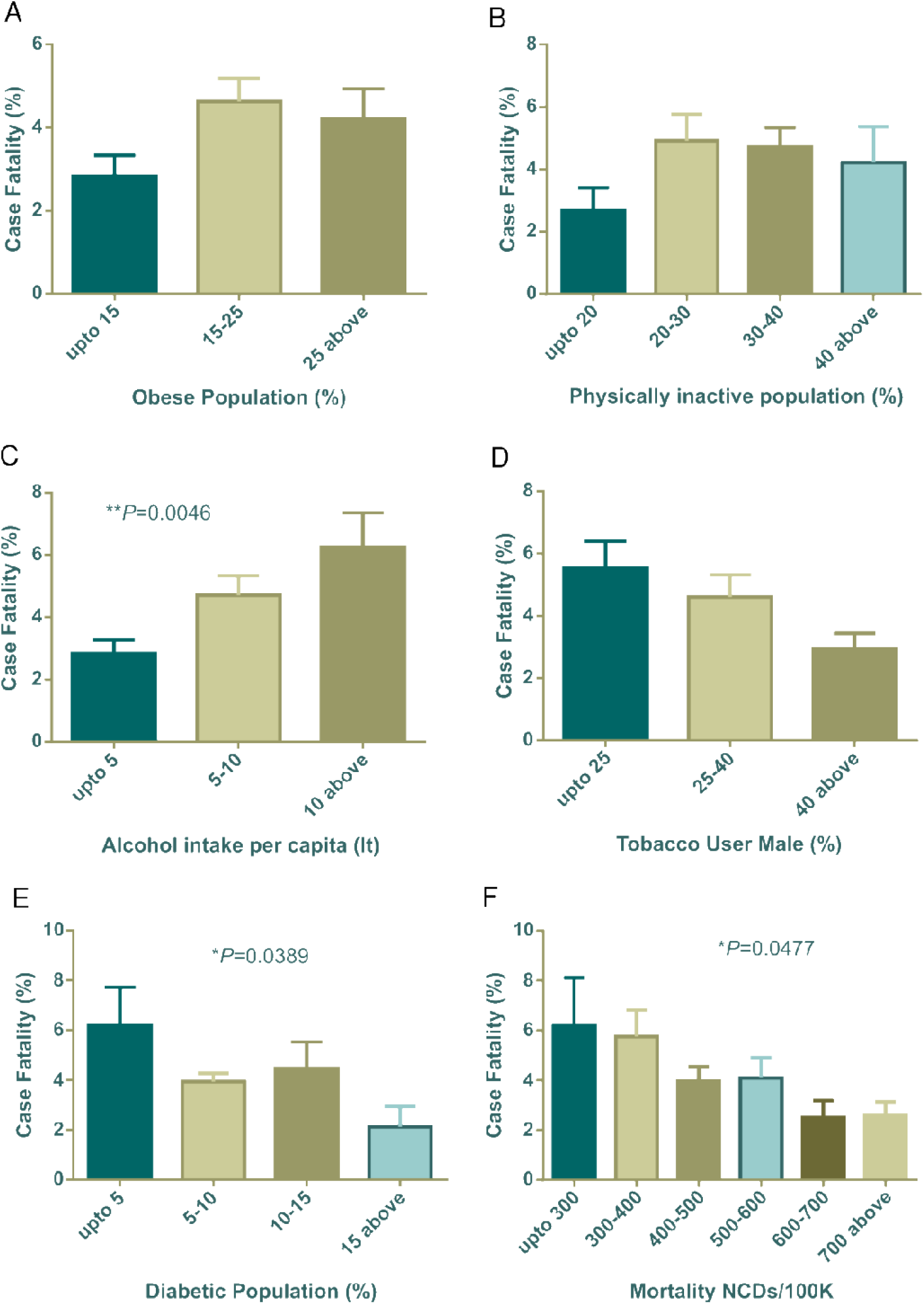
COVID-19 case fatality (%) distribution based on countries grouped according to health indicators (A-F). Alcohol intake, diabetic population and NCDs mortality/100K were statistically significant by one way ANOVA. Data presented are mean ± SEM.

#### 3.5.2 Lifestyle and health indicators on case recovered rate of COVID-19

Our analysis (Figure 5A) showed countries with increasing obesity prevalence were related to the COVID-19 case recovered rate. Insufficient physical activity did not show any meaningful association with recovery rate (Figure 5B). However, high alcohol consumptions displayed relations with high recovery though it was not statistically significant (Figure 5C). Tobacco used pattern did not showed any trends (Figure 5D). Highest prevalence of diabetes exhibited low case recovered rate (Figure 5E) compare to other groups of countries. Low NCD mortalities per 100K groups had relatively high case recovered rate (Figure 5F).

**Figure 5:**
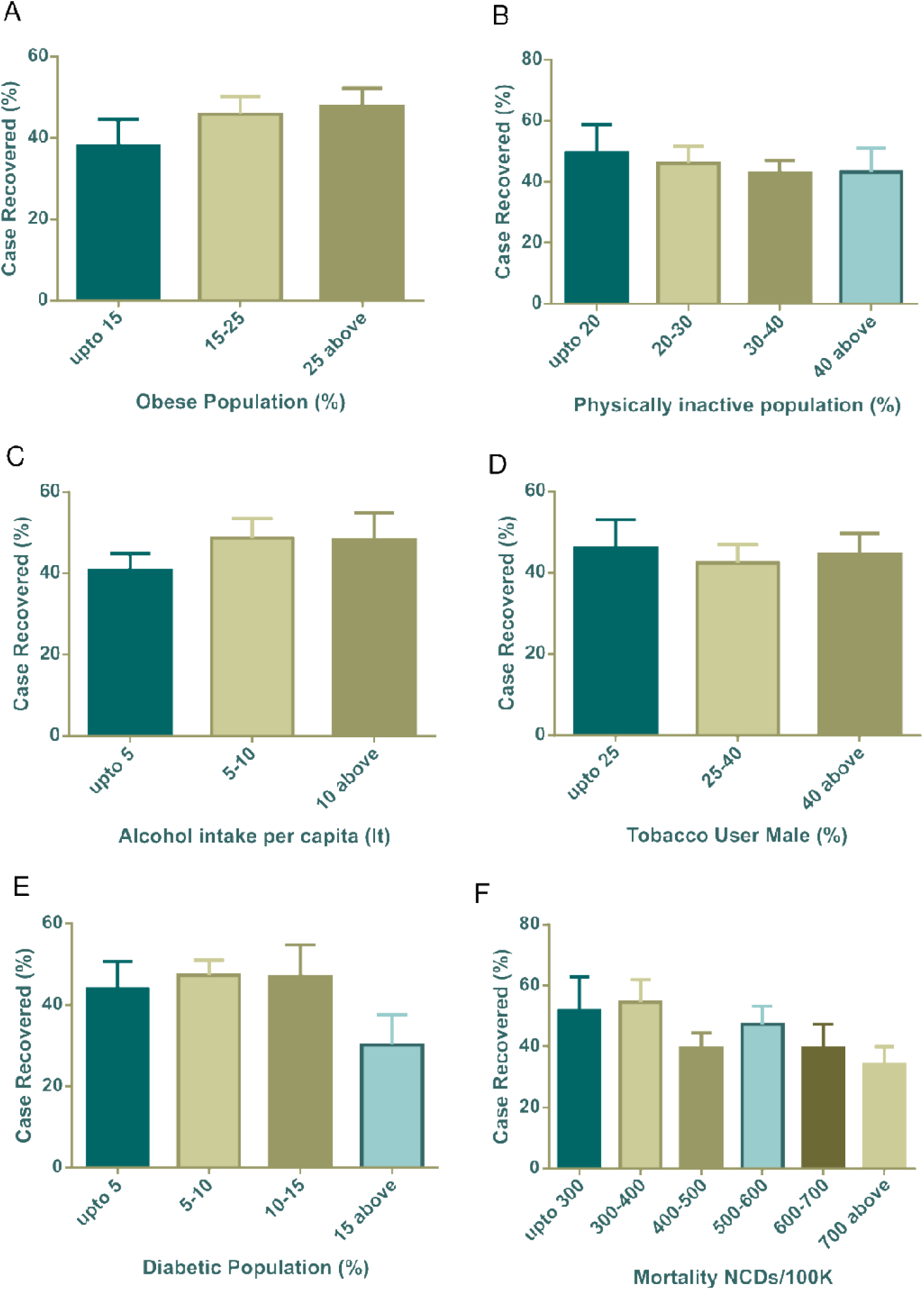
COVID-19 case recovery (%) distribution based on countries grouped according to health indicators (A-F). None of the parameters were statistically significant by one way ANOVA. Data presented are mean ± SEM.

### 3.6 Parametric distribution of countries with top case fatality rates

Association of different factors with dependent variables showed mixed pattern. In this context we further wanted to explore countries with top fatality rates so that any parametric trend can be observed. To do that, we have selected 30 countries with more than 5% case fatalities among 91 countries (**Table 3**). Mean ± SEM values of all parametric distributions are also shown in the Table 03. Individual parameter wise distributions are shown as pie charts in Figure 6 and Figure 7.

**Figure 6:**
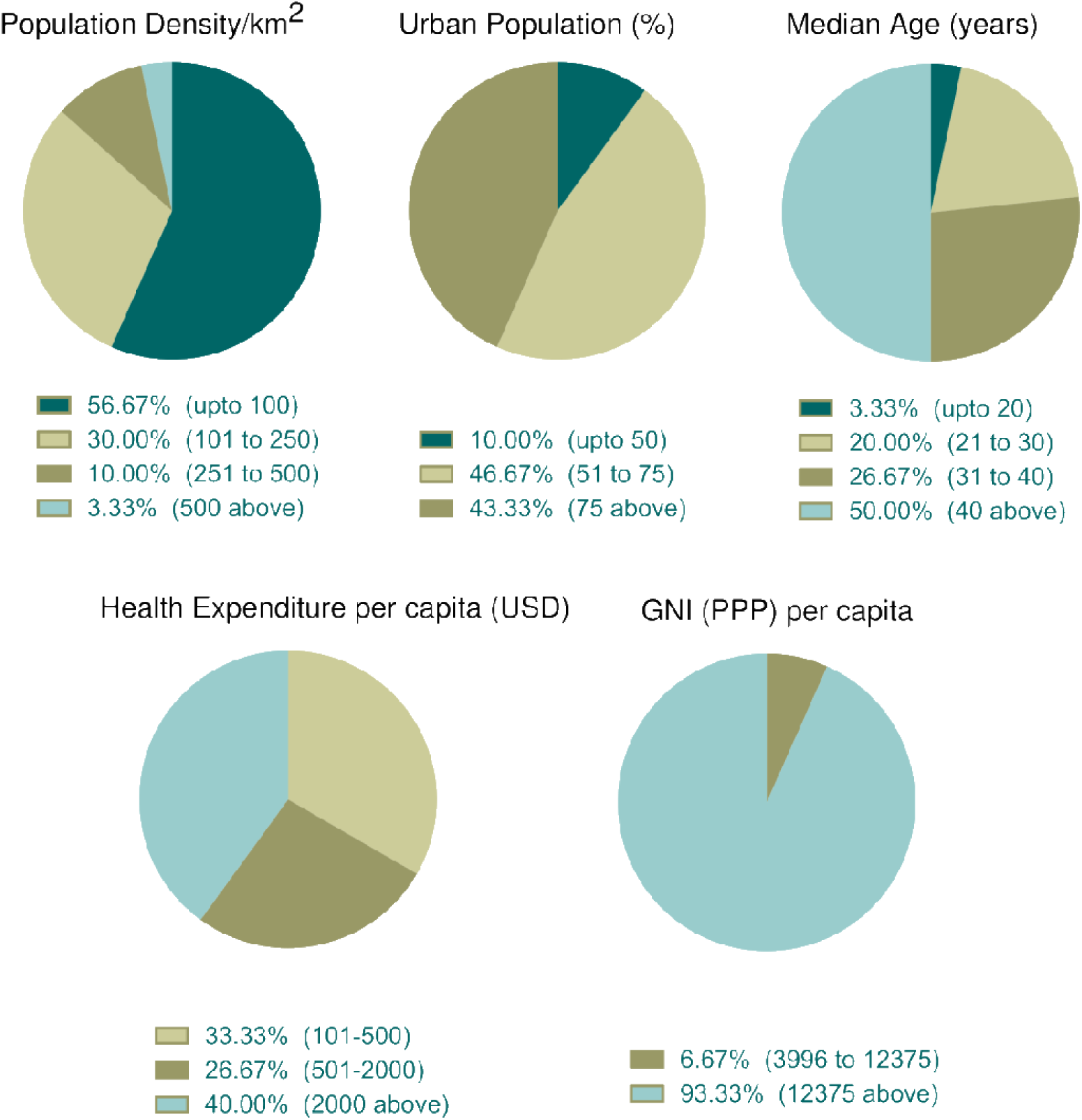
Incidence of socioeconomic and demographic parameter distribution among top thirty countries with high case fatality (%). Data categories with zero data are not shown in the pie chart.

**Figure 7:**
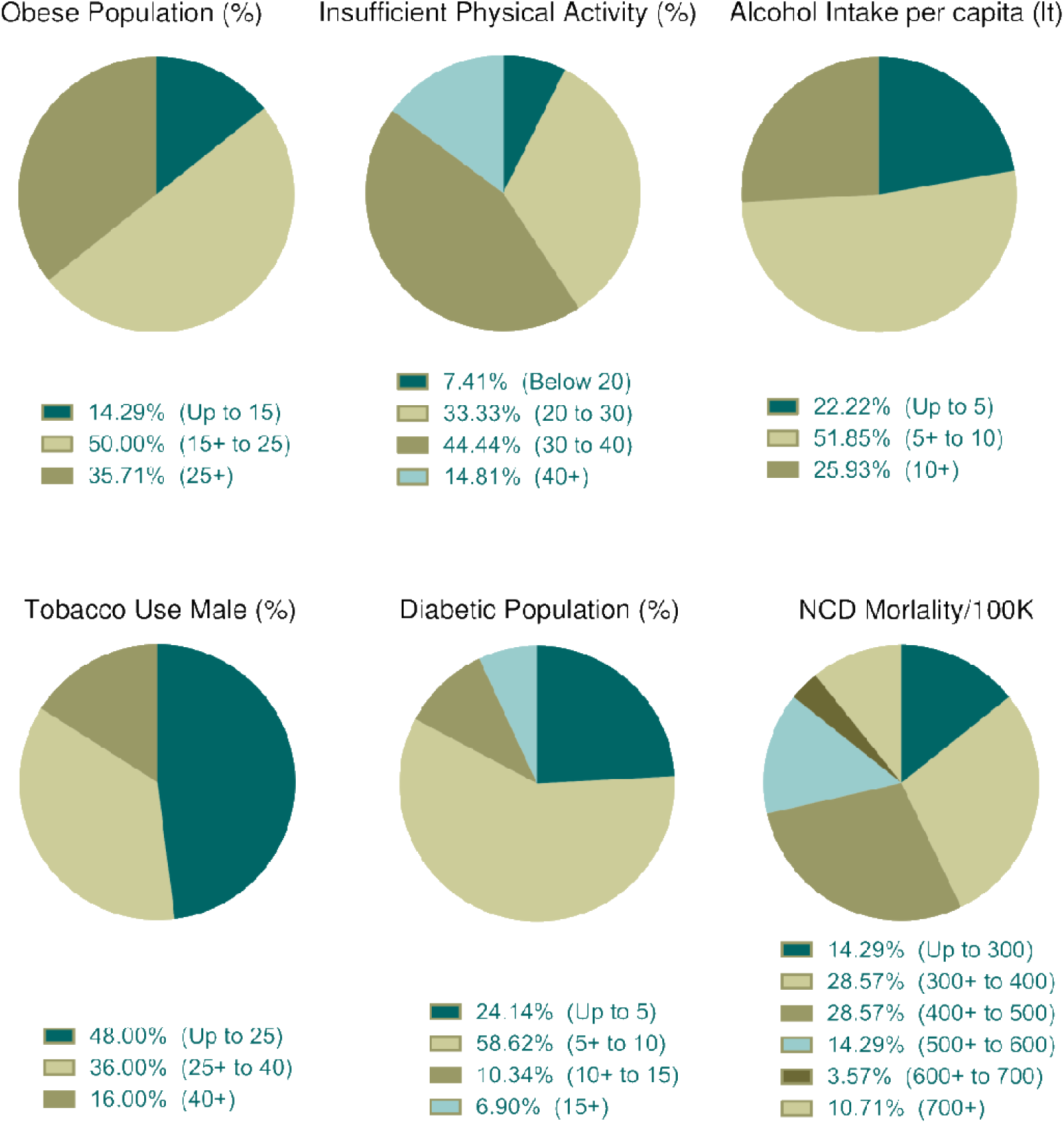
Distribution of health indicators incidence among top thirty countries with high case fatality (%). Data categories with zero data are not shown in the pie chart.

**Table 3:** Top 30 countries according to case fatality rate with their parametric distributions.

| Country | Per capita GNI (PPP),<br>USD | Population Density,<br>KM<sup>2</sup> | Urban Population<br>Percentages | Median age, Years | Per capita health<br>expenditure, USD | Prevalence of Obesity, % | Prevalence of insufficient<br>physical activity, % | NCD Mortality per 100K<br>people | Prevalence of diabetes, % | Alcohol Consumption,<br>per capita in Litter) per<br>year | Prevalence of any kind of<br>tobacco use by male, % |
| --- | --- | --- | --- | --- | --- | --- | --- | --- | --- | --- | --- |
| Sweden | 54030 | 25.00 | 87.43 | 41.10 | 5904.58 | 8.6 | NA | 745 | 22.1 | 0 | 0 |
| Slovenia | 37450 | 102.64 | 54.54 | 44.90 | 1920.28 | 27.4 | 33.6 | 446.6 | 6.7 | 0.6 | 0 |
| Mexico | 19340 | 64.91 | 80.16 | 29.30 | 494.68 | 22.1 | 47 | 452 | 14.2 | 6.5 | 0 |
| Greece | 29670 | 83.22 | 79.06 | 45.30 | 1516.59 | 18.9 | 11.5 | 768 | 5.7 | 9.3 | 0 |
| Denmark | 56410 | 138.07 | 87.87 | 42.00 | 5800.15 | 19.9 | 27.2 | 405.1 | 5.5 | 3.3 | 12.3 |
| USA | 63690 | 35.77 | 82.26 | 38.50 | 10246.14 | 19.7 | 28.5 | 356.6 | 8.3 | 9.5 | 13.4 |
| Brazil | 15850 | 25.06 | 86.57 | 33.20 | 928.80 | 29.4 | 28.6 | 291.6 | 7.6 | 8.1 | 14.8 |
| Algeria | 14970 | 17.73 | 72.63 | 28.90 | 258.49 | 28.9 | 28.9 | 457.8 | 13.5 | 5.5 | 17.3 |
| Philippines | 10740 | 357.69 | 46.91 | 24.10 | 132.90 | 27.8 | 35.9 | 342.4 | 3.9 | 9.8 | 17.3 |
| Canada | 47590 | 4.08 | 81.41 | 41.80 | 4754.95 | 36.2 | 40 | 417.9 | 10.8 | 8.8 | 17.4 |
| North Macedonia | 15670 | 82.59 | 57.96 | 39.00 | 328.42 | NA | 23.1 | 318.3 | 4.8 | 7.2 | 17.5 |
| Hungary | 29860 | 107.91 | 71.35 | 43.60 | 981.42 | 25.8 | 33.2 | 560.8 | 9.6 | 0 | 19.4 |
| France | 46360 | 122.34 | 80.44 | 41.70 | 4379.73 | 25.3 | 32.7 | 348.2 | 3.2 | 11.3 | 20 |
| Ecuador | 11420 | 68.79 | 63.82 | 28.80 | 518.03 | 20.2 | 32.2 | 352.4 | 5.9 | 10.8 | 20.4 |
| Indonesia | 12670 | 147.75 | 55.33 | 31.10 | 114.97 | 22.1 | 35.7 | 333.5 | 4.6 | 10.4 | 24.2 |
| Ireland | 67050 | 70.45 | 63.17 | 37.80 | 4976.86 | 19.5 | 23.7 | 285.4 | 5.7 | 9.5 | 24.7 |
| Belgium | 51740 | 377.21 | 98.00 | 41.60 | 4507.36 | 28.3 | 41.6 | 424.8 | 5.9 | 8.4 | 26 |
| Sudan | 4430 | 21.30 | 34.64 | 18.30 | 193.79 | 19.9 | 41.4 | 306.4 | 5 | 7.1 | 26.9 |
| Honduras | 4790 | 85.69 | 57.10 | 24.40 | 195.94 | 21.6 | 29.3 | 290 | 4.8 | 11.8 | 27.7 |
| United Kingdom | 45350 | 274.83 | 83.40 | 40.60 | 3858.67 | 21.4 | NA | 442.4 | 7.3 | 2.9 | 27.8 |
| China | 18170 | 148.35 | 59.15 | 38.40 | 440.83 | 23.8 | 26.8 | 297.4 | 6.9 | 8.5 | 28 |
| Argentina | 19870 | 16.26 | 91.87 | 32.40 | 1324.60 | 26.4 | 38.5 | 580.2 | 6.9 | 10.9 | 28.3 |
| Switzerland | 68820 | 215.52 | 73.80 | 42.70 | 9956.26 | 23.1 | 32.5 | 467.8 | 6.1 | 10.5 | 28.9 |
| Netherlands | 56890 | 511.46 | 91.49 | 42.80 | 4911.44 | 22.5 | 35.4 | 577.2 | 6.9 | 10.4 | 33.1 |
| Poland | 30010 | 124.04 | 60.06 | 41.90 | 906.82 | 6.4 | 39.7 | 678.3 | 7.1 | 4.5 | 39 |
| Spain | 39800 | 93.53 | 80.32 | 43.90 | 2506.46 | 6.2 | 14.1 | 542.4 | 9.2 | 5.7 | 45.7 |
| Italy | 42290 | 205.45 | 70.44 | 46.50 | 2840.13 | 24.9 | 37.7 | 340 | 4.7 | 6.4 | 49.7 |
| Iran | 21050 | 50.22 | 74.90 | 31.70 | 475.48 | 32 | 31 | 826.7 | 17.2 | 0.2 | 56.3 |
| Egypt | 12100 | 98.87 | 42.70 | 24.10 | 105.77 | 6.9 | 22.6 | NA | 6.3 | 0.3 | 82.7 |
| Romania | 27520 | 84.64 | 54.00 | 42.50 | 555.10 | NA | NA | NA | NA | NA | NA |
| **Mean ± SEM** | **32520.00**<br>**±3573.60** | **125.38**<br>**±21.62** | **70.76**<br>**±2.91** | **36.76**<br>**±1.40** | **2534.52**<br>**±516.47** | **21.97**<br>**±1.41** | **31.57**<br>**±1.56** | **451.97**<br>**±29.21** | **7.81**<br>**±0.78** | **6.83**<br>**±0.71** | **24.79**<br>**±3.31** |

*[Data sources are mentioned in methodology and in supplements. Countries were selected decisively whose case fatality rates were above 5%. NA= Data not available]*

#### 3.6.1 Socioeconomic and demographic distribution among countries with top case fatality rates

Socioeconomic and demographic distributions among top 30 countries whose case fatality rates are more than 5% were illustrated in Figure 6. According to data, more than half of the top 30 countries (n=17, 56.67%) had lowest population density per km^2^. Percentage of urban population was high (above 50%) in 90% countries (n=27). Half of these countries had older population (median age over 40 years). About 40% countries (n=12) spend more money in health sectors per capita and 93.33 % (n=28) countries belong to high income group.

#### 3.6.2 Lifestyle and health parameter distribution among countries with top case fatality rates

Prevalence of obesity was observed in 28 countries. Among them 50% countries belong to moderate percentage of obese population (Figure 7). Prevalence of insufficient physical activities data were available for 27 countries and among them 44.44% (n=12) countries whose 30% to 40% people were physically inactive (Figure 7). About 51.85% (n=14) countries among 27 consumed 5 to 10 liters of alcohol per capita per year and nearly half of these countries (n=12 of 25, 48%) were used less tobacco products (Figure 7). About 58.62% countries (n=27 of 29) were moderately diabetic (5% to 10% diabetic prevalence) and more than half (n=16 of 28, 57.14%) of the countries were at risk of death whose NCDs mortality rate per 100K population was low to moderate (Figure 7).

## 4 Discussion

### 4.1 Socioeconomic and demographic determinants of COVID-19 test positive, case fatality and case recovery rate

Population density both in a country and in urban cities can have impact on contagious disease transmission. Both contact rates and pattern in a defined geographical spatial distribution determine the mode of infectious disease transmission (47, 48). Contact hours and contact number per person vary with age and number of household members. In a study in Hong Kong the highest contact pattern was observed among school going children, which decreased with age (48). However, older age group with economic strength also showed elevated contact rates. We found weak relation of population density per km^2^ with percentage of test positives (Figure 1A). Generally, population density poses increased chance of disease transmission, but contact rate is not always determined by population density. At low density contact rate can increase rapidly which gets saturated at very dense area where lack of organized social contact is evident (47). At this point contact rate becomes independent of population density. Although in our analysis low density countries showed slightly higher percentage of test positives, further data analysis showed that few countries in this group e.g. Algeria, Brazil and Afghanistan had very high test positive value as outliers as they did very low number of testing. This inadequate testing increased their test positive rate as we found in our analysis too (Figure 3B).

Number of tests performed to confirm COVID-19 is still a challenging issue for many countries and it can affect decision making drastically. We found statistical linear positive relation in number of positive patients with increasing test numbers (Figure 3A). The following graph depicted that as the number of tests per 100K population increased, percentage of test positives dropped gradually (Figure 3B). In a recent press conference WHO mentioned that high percentage of test positive in a country indicate inadequate number of tests performed and they set a rough standard of 10% test positives as indicator of normal outcome (49). This strongly supports our data in Figure 3B.

Countries that performed above 500 tests per 100K population, showed around 10% test positives. Lower middle and upper middle economy countries performed symptom-based highly selective diagnosis due to socioeconomic, demographic and political reasons which increased their percentage of test positives.

Test positive rate also has close link with urban population, median age and income level. Among 91 countries in our study, around 74.73% (n=68) countries belong to high economy class, 20.88% (n=19) were from upper middle economy and rest were from lower middle economy group (Table 1). Thus, the most of the independent socioeconomic and demographic variables comply with the characteristics of developed countries. These countries possess modern health facilities, medical professionals and cutting-edge research facilities. Despite having these benefits in health sectors they could not successfully restrict infections and deaths toll. However, these amenities certainly were advantageous in some form and contributed in patient recovery (Figure 2D).

Developed countries own bigger number of city populations with a big proportion of older inhabitants who prefer to stay at home (50, 51). The test positive rates thus were lower in groups of countries with dense urban populations (Figure 1B). Median age data presented that developing countries hold more young populations compared to developed countries. Young population always have tendencies to go outside and making them vulnerable age group (32). Therefore, test positive rates were high in developing countries where median age is below 30 years (Figure 1C). Health expenditure per capita correlates with high number of tests performed, thus countries with over 501 USD investments showed decreased test positive rate (Figure 1D). As high median age belongs to countries with strong healthcare system, case recovery was also found to be high where median age is over 30 years (Figure 2C)

Case fatality and recovery from epidemics largely depends on a person’s internal and external factors along with age, presence of co-morbidities, health facilities and pattern of adherences at healthcare centers (25, 33, 50, 52). The effect of contagions on certain population is also influenced by the interplay between incubation period and the age dependent case fatality rate of the disease (53). In our study, case fatality rate was high where median age is over 40 years (Figure 1C) and in countries with more than 50% urban population (Figure 1B). Although increasing population density showed gradual rise in case fatality, countries with over 500 people per km^2^ density (n=6) surprisingly showed reduced case fatality (Figure 1A). Most of the countries in this group have high urban population and high median age; however, they managed to control case fatality with remarkable success, except the Netherlands with 12.79% case fatality. With rapid urbanization the risk of pandemic and zoonotic diseases are increasing (54). Being infectious and contagious disease, urban population percentages is one of the important predictors in COVID-19 outcome. In countries with more people living in rural areas have low case fatality as air velocity has potential reducing effect on disease transmission (55). In contrast, urban medical facilities, availabilities of required medicines/drugs, and treatment tools accelerate disease recovery despite huge load of patients compared to rural and remote areas.

Countries with high health expenditure per capita and GNI (PPP) could not restrict the pace of death as expected due to inadequate intensive care equipment and management personnel with overwhelming number of critical patients. Although, better urban intelligence and management is partially linked with better recovery (56), as proportion of urban population around the globe is gradually increasing, policy level rethink on healthcare system is a must. We need to redesign urban intelligence, resource management and coordination to fight COVID-19 like pandemic where isolation, social distancing and quarantine are challenging (57).

Investment on health specially toward NDCs have shown reduced mortality/incidence ratio in cancer (58), stoke (59) and child mortality (60). Expenditure to contain infectious disease is still a necessary component; as controlled use of available resources can reduce disease spreading (61). Moreover, sudden pandemic induced resource constraints can critically affect treatment and patient recovery (62). Contrary to misconceptions that higher expenditure relate to better healthcare, developed countries showed no strong negative correlation of health expenditure to case fatality (63). This could be due to inappropriate way of spending money, poorly designed policy and political intrusions that inhibit policy and treaty implementation (64). So far in COVID-19 pandemic, some countries offered strict preventive measures and information technology based contact tracing to successfully contain the disease although they fall into high risk group countries. In our study, COVID-19 related case fatality rate was comparatively higher in countries with high health expenditure per capita which explains that majority of health investment is insufficient or inadequate in required segments to tackle such pandemic. In a recent study, COVID-19 incidence and case fatality was found not to be associated with health expenditure and services (34). The situation gets even worse as incidence number rise rapidly putting pressure on healthcare capacity, as observed in some European countries (25).

### 4.2 Lifestyle and health indicators on case fatality rate

Duration of infectious disease is very important and had direct impact on mortality (53). Clinical recovery from COVID-19 cases requires approximately 14 to 42 days depending upon patient’s physical and clinical conditions (65). Thus, unhealthy lifestyles are important predictors of delayed recovery or death. Lifestyle includes obesity, physical activities, pattern of drinking alcohol, tobacco usages etc. Besides, this prevalence of NCDs and their mortality also have linked to the raised number of case fatality rate.

Several factors including obesity, diabetes, cardiovascular disease and hypertension, cancer, chronic respiratory diseases, have been identified as collective underlying conditions of critical illness and poor outcomes in COVID-19 (66, 67). Recent correspondences indicated that obesity and insufficient physical activity might accelerate the mortality of COVID-19 (66, 67). In our study, countries with large number of obese and insufficiently active populations behold the increased number of case fatality rates. This result complies with the correspondences statement. However, Center for Disease Prevention and Control (CDC), USA, listed severe obesity (BMI over 40) as risk factors for critical COVID-19 cases (68). Furthermore, clinical reports of critical COVID-19 patients showed that, significant numbers of patients are associated with obesity in different countries (66). Obesity and insufficient physical activity lead to metabolic, cardiovascular disorders and other NCDs (69).

Though the relationships between obesity and NCDs are well studied, a little is known about the effect of obesity on immunity and contagious disease. A recent clinical studies on survivors and non-survivors of COVID-19 showed that inappropriate and abnormal immunity were significantly associated with death (70). Several animal model studies showed that obesity leads to impairment of natural killer cell, reduction of macrophages and dendritic cell activities (69), with reduced cytokine productions and weakened responses to antigen stimulations. Thus, impaired immune systems cannot fight the pathogen resulting delayed recovery or death.

Studies showed that diabetes is the second top co-morbidity factor that influences the COVID-19 case fatality after hypertension (66). Surprisingly, our analysis demonstrated that, group of countries with lowest prevalence of diabetes observed more case fatality (Figure 4E). To explain this outcome, we further analyzed group prevalence of diabetes with group median age as diabetes is more prone to elderly people (supplementary Table 1). The analysis demonstrated that 55.56% (n=10 of 18) countries holding up to 5% diabetes prevalence comprises the median age over 40 years and none of the countries with 15% diabetes prevalence were from the same median age group of countries. Further cross analysis with GNI (PPP) per capita (supplementary Table 2) confirmed that 72.22% developed countries were from the group of less prevalence of diabetes. This result established that diabetes is one of the key factors on case fatalities as earlier we have demonstrated that case fatalities were more evident in elderly people and in the developed countries.

In addition, diabetes is characterized by the chronic elevated levels of blood glucose. This raised blood sugar concentrations also increase the glucose concentration in airway secretions (71). A study demonstrated that in vivo influenza virus infection and replications can be significantly increased due to the contact of pulmonary epithelial cells with raised glucose concentrations in vitro (72). In addition, raised sugar levels in blood perhaps weakened the anti-viral immune response and can be reverted with insulin treatment (73). Another studies showed that high glucose concentration or diabetic conditions were associated with fatal outcome of avian influenza (74). Therefore, it could be concluded that, chronic diabetic conditions can elevate case fatality rate of COVID-19.

Countries with less NCDs mortality per 100K population faced more COVID-19 death rates (Figure 4F). To explain this, we again did cross analysis of group NCDs mortality per 100K with group median age and group GNI (PPP) per capita. Around 71.43% of the countries of lowest NCD mortality per 100K belong to the highest group of median age and all of them were high income group countries (supplementary Table 3 and Table 4). As discussed above, case fatalities were high in developed countries and the cross analysis of NCDs mortality with median age and per capita income also suggested that elderly people with NCDs were more risk of fatal outcome. Most of the mortality (60% to 90%) was related to preexisting one or more NCDs (75-78).

Heavy alcohol intake has been casually associated with several disease including infectious disease (79). This could be explained as heavy intake of alcohol weakened the immune function as well as several organs including liver and lung and make susceptible to microorganisms (79) and substantially lowers the adherence to the antiretroviral therapy accelerating the mortality (80). In our investigations, it was showed that case fatality was increased with per capita alcohol intake in liters (Figure 4C).

Tobacco smoking is generally linked with lung diseases (81) and can facilitate microbial infection (82). There is a scientific debate going on about the relation between tobacco smoking and COVID-19 severity (83). A recent systematic review concluded that the smoking might be negatively linked to the COVID-19 case fatality (84). However, another short meta-analysis stated no association between them (85). In addition, a group of French scientists showed that tobacco smoking has negative effects of COVID-19 mortality (86) which was also reflected in our results (Figure 4D). The recent news (87, 88) reported that French scientists are planning to a human trial to test that tobacco can fight against COVID-19. As the debate is going on, detail clinical and molecular investigation is required to find the right answer.

### 4.3 Limitation and future direction

COVID-19 pandemic is ongoing and not yet closed; data used in our study reflects a snapshot of a time point. This limits our study to capture full view of the dynamic nature of this disease. In addition, self-reported government data often pose reliability issues. As COVID-19 is multifactor mediated, not all factors could be included in this particular study specially integrating molecular mechanism of disease was beyond the scope. In addition, as molecular mechanism of pathogenesis will gradually unfold, and more clinical data will be available, the factors discussed in this study will be easier to interpret. Finally, the interpretation presented in our study can be useful to design future plans to contain such contagious pandemic outbreak within very short time.

## Data Availability

All secondary data sources are mentioned in manuscript and readily available for researchers.

## 6 Author Contributions

AA and MMR conceived and designed the study. MMR and TH collected and sorted the data from different sources. AA and MMR conducted the analysis with input from TH. AA and MMR equally contributed to the first draft and TH added additional points in discussions. After necessary corrections and suggestions from all authors AA finalized the draft and agreed to submit the manuscripts.

## 8 Acknowledgments

This is a short text to acknowledge the contributions of specific colleagues, institutions, or agencies that aided the efforts of the authors.

